# Persistent humoral immunity in children and adolescents throughout the COVID-19 pandemic (June 2020 to July 2022): a prospective school-based cohort study (Ciao Corona) in Switzerland

**DOI:** 10.1101/2023.05.08.23289517

**Authors:** Alessia Raineri, Thomas Radtke, Sonja Rueegg, Sarah R. Haile, Dominik Menges, Tala Ballouz, Agne Ulyte, Jan Fehr, Daniel L. Cornejo, Giuseppe Pantaleo, Céline Pellaton, Craig Fenwick, Milo A. Puhan, Susi Kriemler

## Abstract

**Objectives:** To assess the longitudinal development of humoral immunity in children and adolescents during the COVID-19 pandemic, with a particular focus on how anti-spike IgG antibodies and neutralising response changed during the first Omicron peak (December 2021 to May 2022).

**Design:** Prospective school-based study during the COVID-19 pandemic (June 2020 to July 2022) including five testing rounds with corresponding cross-sectional cohorts and a longitudinal cohort who participated in at least four rounds.

**Setting:** 55 randomly selected schools in the Canton of Zurich, Switzerland.

**Participants:** Between 1875 to 2500 children and adolescents per testing round and 751 in the longitudinal cohort.

**Main outcome measures:** Development of SARS-CoV-2 seroprevalence, anti-spike IgG antibodies and neutralising antibody response over time, persistence of antibodies and variation of antibody levels in individuals only infected, vaccinated or with hybrid immunity during the early Omicron period.

**Results:** By July 2022 96.9% (95% credible interval [CrI] 95.2 to 98.1%) of children and adolescents had anti-spike IgG antibodies against SARS-CoV-2. The substantial increase in seroprevalence during the first peak of the Omicron wave was largely driven by primary infections in mostly unvaccinated children under the age of 12 (28.4% [95% CrI 24.2 to 33.2%] in December 2021, to 95.7% [95% CrI 93.4 to 97.4%] in July 2022). This stands in contrast to adolescents aged 12 years and older (69.4% [95% CrI 64.0 to 75.4%] in December 2021 to 98.4% [95% CrI 97.3 to 99.2%] in July 2022), who were eligible for vaccination since June 2021. Children and adolescents with hybrid immunity or immunity from vaccination had high anti-spike IgG titres (median Mean Fluorescence Intensity (MFI) ratio of 136.2 [Inter Quartile Range [IQR]: 121.9 to 154.3] and 127.6 [IQR: 114.1 to 151.0]) and strong neutralising responses (e.g., anti-Omicron 98.9% [95% Confidence Interval [CI] 96.0 to 99.7%] and 81.6% [95% CI 74.9 to 86.9%]). Meanwhile, infected but unvaccinated children and adolescents had substantially lower anti-spike IgG titres (median MFI ratio of 54.8 [IQR: 22.8 to 89.8]) and neutralising responses (e.g., anti-Omicron 64.9% [95% CI 59.8 to 69.7%]).

**Conclusion:** These findings show that the Omicron wave and the rollout of vaccines led to almost 100% seropositivity and boosted anti-spike IgG titres and neutralising capacity in children and adolescents. This was particularly driven by unvaccinated children (<12 years), who became seropositive due to the highly infectious Omicron variant. Nevertheless, during the entire study period parents of only one adolescent reported hospital stay of less than 24 hours related to a possible acute infection.

## Introduction

Monitoring the evolution of seroprevalence and assessing changes in humoral immunity against severe acute respiratory syndrome coronavirus type 2 (SARS-CoV-2) in children and adolescents over time is important to understand how the pandemic evolved and to inform public health measures including vaccination strategies as well as preventive measures at school.

Several serological studies were conducted to detect SARS-CoV-2 infections in children and adolescents and to determine seroprevalence at different times of the pandemic [1–6]. However, little is known about the development and persistence of those antibodies over time as most studies were cross-sectional [1–5] and there is only limited data from longitudinal studies assessing the humoral immunity in children and adolescents [7]. A systematic review [7] reported persistence of cellular and humoral immunity in the pre-Omicron period lasting for at least 10 to 12 months in children and adolescents. Meanwhile, few studies [8, 9] focused on immune responses following Omicron, addressing neutralising activity and differentiating between natural infection, vaccination, or both [10].

Many countries started to administer COVID-19 vaccines to children and adolescents in 2021 to 2022, after trials demonstrated the effectiveness of the COVID-19 vaccine against re-infection [11, 12] or severe disease [13], and vaccination was approved by the Food and Drug Administration and European Medicines Agency. In Switzerland, the COVID-19 vaccine was available for adolescents aged 12 years and older by mid 2021 and for children aged 5 to 11 years in early 2022 [14].

In the beginning of 2022, the high incidence of SARS-CoV-2 infections in children and adolescents due to the highly infectious Omicron variant despite the rollout of vaccines raised concerns [15–17]. In that period, the coincidence of incomplete immunisation of youth and the highly transmissible Omicron variant affected the evolution of seroprevalence and the longitudinal development of the humoral immunity in children and adolescents.

In this observational school-based study, we aimed to assess the longitudinal development of the humoral immunity in children and adolescents during the COVID-19 pandemic with a particular focus on how anti-spike IgG antibodies and neutralising response changed during the first Omicron peak in the context of (re-)infections, vaccinations, or their combination.

## Method

### Study setting & design

The Ciao Corona study is embedded in a nationally coordinated research network *Corona Immunitas* in Switzerland [18]. The protocol of the study was registered prospectively (ClinicalTrials.gov identifier: NCT04448717) [19], and seroprevalence results of the first four Ciao Corona testing rounds can be found elsewhere [20–23]. This repeated cross-sectional analysis is based on a prospective cohort study, using data from children and adolescents who participated at multiple timepoints. The study took place in the canton of Zurich with its around 1.52 million (18% of the Swiss population) ethnically and linguistically diverse inhabitants and comprises rural as well as urban regions.

In March 2020, the first restrictions and preventive measures were announced from the Swiss Federal Office of Public Health. Schools closed on 16 March 2020 and partially reopened on 10 May 2020, with a combination of in-person and online teaching. On 7 June 2020, schools resumed usual in-person teaching with certain preventive measures (e.g., contact tracing systems within schools, mandatory face mask for school personnel, distancing regulation). Implementation of restrictions varied across schools. For adolescents of 12 years or older, masks were mandatory starting from October 2020 and for children between 9 to 11 years starting from January 2021. This was implemented due to an increase in the incidence of SARS-CoV-2 infections, signaling a second pandemic wave. Throughout summer 2021, masks were no longer mandatory for children and adolescents. However, they were reinstated for all school children and adolescents from December 2021 to mid-February 2022 during the first peak of the Omicron wave. Adolescents of 16 years or older were allowed to get vaccinated starting from May 2021, adolescents between 12-15 years of age since mid-June 2021 and children between 5 and 11 years of age from January 2022 [14].

### Population

We randomly selected primary schools in the canton of Zurich and invited for each primary school the secondary school that was the closest geographically. The number of invited schools per district corresponded to the population size of the 12 districts. Out of 156 invited schools, both public and private (around 10%), 55 schools agreed to participate. Classes were randomly selected and stratified by school level: grades 1-2 (6 to 8 years old children) of lower school level, grades 4-5 (9 to 11 years old children) of middle school level, and grades 7-9 (12 to 14 years old adolescents) of upper school level. All children and adolescents in the randomly selected classes were eligible to participate in any of the testing rounds, irrespective of whether they participated at baseline.

### Timeline of testing

Venous blood samples were collected in five testing rounds. The first testing round (T1) was performed in June/July 2020, the second (T2) in October/November 2020, the third (T3) in March/April 2021, the fourth (T4) in November/December 2021 and the last fifth (T5) testing round in June/July 2022. As shown in the study participant flow chart (Figure 1), we followed corresponding repeated cross-sectional cohorts and a longitudinal cohort. The longitudinal cohort consisted of children and adolescents participating in the last (T5) and at least three previous testing rounds. In addition, we only included children and adolescents where the seroconversion from one to the subsequent testing round was detectable.

**Figure 1:**
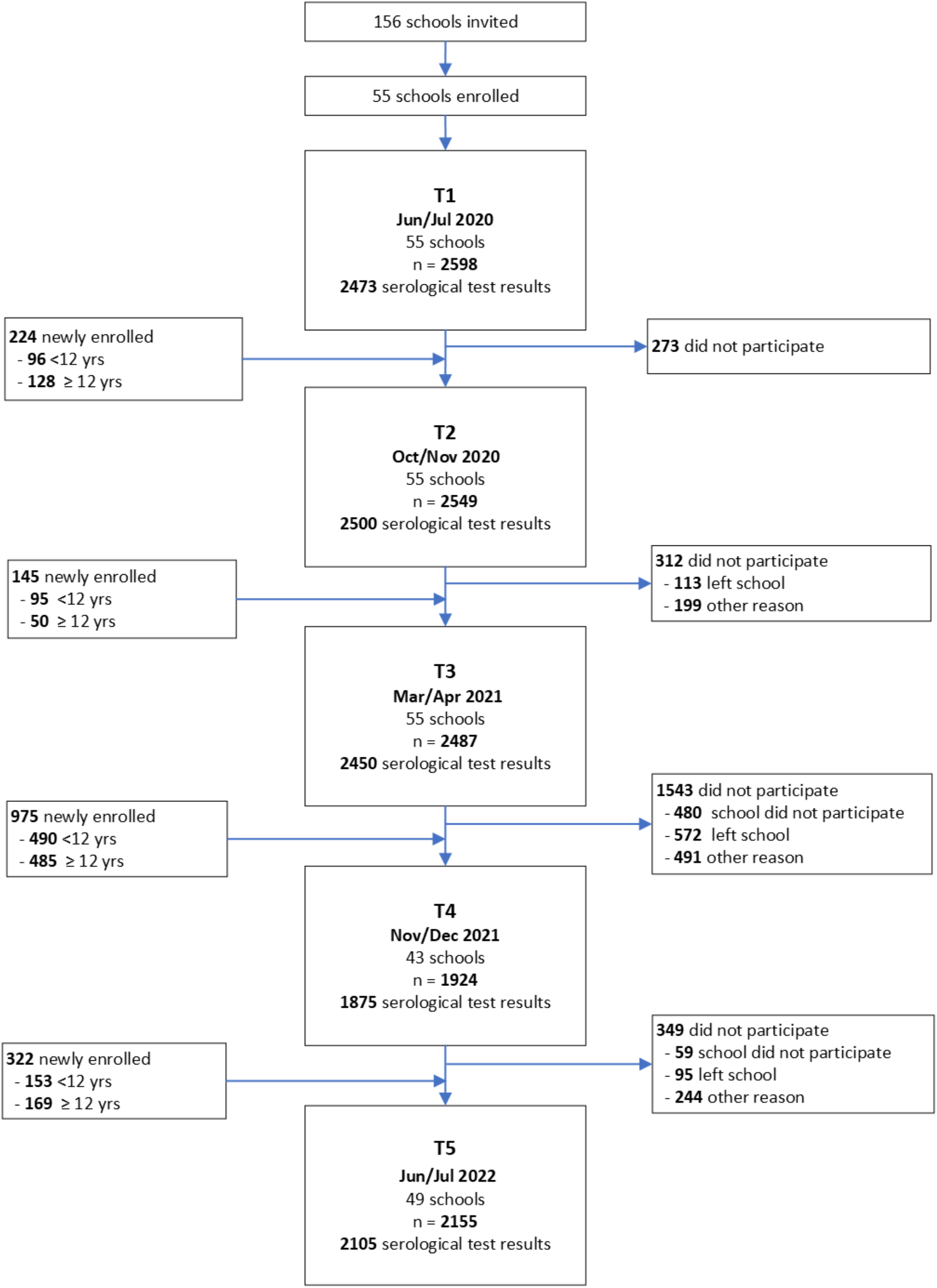
*Flowchart of participants. Newly enrolled children and adolescents did not participate in previous rounds. *Longitudinal cohort consists of children and adolescents participating in the last (T5) and at least three other testing rounds. Yrs: years.*

### Serological testing and neutralisation assay

In all five testing rounds, we visited children and adolescents in schools and collected venous blood samples. To detect SARS-CoV-2 specific antibodies against spike and nucleocapsid proteins we used the Sensitive Anti-SARS-CoV-2 Spike Trimer Immunoglobulin Serological (SenASTrIS) test [24]. This Luminex assay measures the binding of IgG antibodies to the trimeric SARS-CoV-2-spike and nucleocapsid proteins, obtaining the Mean Fluorescence Intensity (MFI) ratio. Test results were considered seropositive if MFI ratios were equal to or above the cutoff of 6 for both anti-spike IgG and anti-nucleocapsid IgG, based on which the test has a 98% specificity and 99% sensitivity [24]. Test validation was performed in different cohorts of pre-pandemic plasma of adults and children and SARS-CoV-2 infected people [24]. We used a cell-free and virus-free assay to detect SARS-CoV-2 neutralising antibodies against the Wildtype SARS-CoV-2, Delta, and Omicron variants, by measuring the proportion of antibodies preventing the binding of the angiotensin-converting enzyme 2 receptor to the receptor binding domain of the trimer spike protein of the different SARS-CoV-2 variants [25]. Neutralising activity was quantified by the half maximal inhibitory concentration (IC50), defining values of 50 or higher as positive [25].

### Questionnaire

Online questionnaires were sent to participants at enrolment and repeatedly every 3 to 6 months over the duration of the study, collecting information on sociodemographic characteristics, chronic conditions, and vaccination status. Vaccination status of children and adolescents was either self-reported by children and adolescent on the day of testing in schools or reported by parents/caregivers in online questionnaires.

### Groups of children and adolescent according to seropositivity and exposure status

To assess the evolution of anti-spike IgG and neutralising antibody titres, we divided children and adolescents from the longitudinal cohort into four groups according to their vaccination and infection status. Children and adolescent never testing positive for anti-spike IgG were categorised as *seronegative*, unvaccinated children and adolescent ever testing positive for anti-spike IgG as *infected*, vaccinated children and adolescent testing negative for anti-spike IgG prior to vaccination and never testing positive for anti-nucleocapsid IgG as *vaccinated*, and children and adolescents testing seropositive before getting vaccinated, or were vaccinated and tested positive for anti-nucleocapsid-IgG antibodies as *hybrid*.

### Statistical Analysis

We performed descriptive analysis for participants’ characteristics and antibody titres, by reporting median (interquartile range) or count (percentage). Neutralising activity was visualised using a log10-transformation of scales. The Wilson method was used to calculate 95% confidence intervals (95% CI) of proportions [26]. We divided the study population into children being younger than 12 years and adolescents of 12 years and older, based on different vaccination policies for younger and older children and adolescents in Switzerland [14].

We used Bayesian logistic regression to estimate the seroprevalence with 95% credible intervals (95% CrI), using a model which accounts for the sensitivity and specificity of the SARS-CoV-2 antibody test and the cohort’s hierarchical structure. The Bayesian approach also allowed to adjust for geographic district of the school, sex, and school grade of the child, and included random effects for school levels (lower, upper, and middle). We used poststratification weights to adjust for the population size of the particular school level and the geographic district. Further details regarding the Bayesian model and weighting approach can be found elsewhere [20].

Seroprevalence in the first three rounds (T1 to T3) was only referring to unvaccinated children and adolescents since vaccination was only available since June 2021 (rounds T4-T5, see Figure 1). For T4 and T5, we conducted the analysis of seroprevalence for two groups: a) the unvaccinated children and adolescents, and b) all participating children and adolescents.

To determine anti-spike IgG antibody decay times, we included all participants that seroconverted at any testing round and of whom at least one follow-up serology was performed. We excluded a) individuals who never tested seropositive for anti-spike IgG antibody, b) who had no follow-up assessment after testing seropositive, c) those who were vaccinated and d) those with potential reinfection, defined by the presence of anti-nucleocapsid IgG or any increase in anti-spike IgG titres between two testing points. To estimate the slope of antibody decay, we limited the data to the first seropositive result (the closest and therefore likely highest MFI ratio after an infection) and all following timepoints, and then realigned the time axis to begin at the first seropositive result for each individual as done by others [27–29]. We then fit the univariable mixed-effects linear decay model for the natural logarithm of the titres, with random intercepts for each participant. We used the formula (ln(0.5)/ β) to estimate the half-life (λ) in days, with β being the coefficient for time deriving from the fitted model [27–29]. In the primary analysis, we estimated the anti-spike IgG half-life in children and adolescents considering a time window of 365 days. Additionally, we performed a sensitivity analysis, estimating the half-life over a time window of 220 days, to ensure comparability with other published studies [8, 29–33]. Compared to other studies that used even shorter time windows for estimating the half-life, we chose a time window of 220 days due to the timing of our testing rounds.

The analyses were performed with R programming language [34], including the RSTAN package to fit the Bayesian models [35].

## Results

### 1. Participant Characteristics

Throughout the study, we tested between 1876 and 2500 children and adolescents at each testing round between June 2020 and July 2022. Figure 1 shows the flowchart of participating schools and children and adolescents, and valid serological test results at each testing round. Table 1 presents the baseline characteristics of the study population for the repeated cross-sectional as well as for the longitudinal cohort (Supplementary Table 1 for more details on chronic conditions). During the entire study period, three seropositive tested children and adolescents reported hospital stays of less than 24 hours, of which one was possibly related to an acute SARS-CoV-2 infection.

**Table 1:** *Baseline characteristics of the study population at each testing round. Yrs: years; a: Predominant variant of concern in Switzerland (>50% of circulating VOC in Switzerland); b) unique number of children and adolescents tested throughout the entire study period; c) number of children and adolescents tested per round; d: details on chronic conditions can be found in Supplement Table 1; e: grouped into <12 years and ≥12 years, since in Switzerland, adolescents ≥12 years of age could get vaccinated since mid-June 2021 and children between 5 and 11 years of age from January 2022; *median (interquartile range)*

|  | T1 | T2 | T3 | T4 | T5 |
| --- | --- | --- | --- | --- | --- |
| **Timeframe of testing** | Jun/Jul 2020 | Oct/Nov 2020 | Mar/Apr 2021 | Nov/Dec 2021 | Jun/Jul 2022 |
| **Predominant VOC <sup>a</sup>** | Wildtype | Wildtype | Alpha | Delta | Omicron |
| **REPEATED CROSS-SECTIONAL COHORTS** |  |  |  |  |  |
| **N unique <sup>b</sup>** | 4251 |  |  |  |  |
| **N tested <sup>c</sup>** | 2473 | 2500 | 2450 | 1875 | 2105 |
| **Age\* (yrs)** | 12 (6-17) | 12 (7-17) | 12 (7-17) | 12 (7-17) | 12 (7-18) |
| **Sex (n, % male)** | 1197 (48%) | 1211 (48%) | 1165 (48%) | 884 (47%) | 990 (47%) |
| **Age group** |  |  |  |  |  |
| <12yr | 1450 (59%) | 1298 (52%) | 1144 (47%) | 945 (50%) | 1011 (48%) |
| ≥12yr | 1023 (41%) | 1202 (48%) | 1306 (53%) | 930 (50%) | 1094 (52%) |
| **Existence of any chronic condition <sup>d</sup>** | 546 (24%) | 551 (24%) | 536 (24%) | 375 (23%) | 412 (23%) |
| **Vaccinated <sup>e</sup>** |  |  |  |  |  |
| Overall | 0 | 0 | 0 | 475/1876 (25%) | 913/2105 (43%) |
| <12yr |  |  |  | 0 | 283/1011 (28%) |
| ≥12yr |  |  |  | 475/930 (51%) | 630/1094 (58%) |
| **Questionnaires completed** | 2211 (89%) | 2030 (81%) | 1898 (77%) | 1461 (78%) | 1499 (71%) |
| **LONGITUDINAL COHORT** |  |  |  |  |  |
| **N unique <sup>b</sup>** | 751 |  |  |  |  |
| **N tested <sup>c</sup>** | 695 | 725 | 738 | 722 | 751 |
| **Age\* (yrs)** | 10 (6-15) | 11 (7-16) | 11 (7-16) | 12 (8-17) | 12 (8-17) |
| **Sex (n, % male)** | 326 (43%) | 341 (45%) | 352 (47%) | 341 (45%) | 355 (47%) |
| **School level** |  |  |  |  |  |
| **<12yr** | 496 (71%) | 492 (68%) | 465 (63%) | 377 (52%) | 325 (43%) |
| **≥12yr** | 199 (29%) | 233 (32%) | 273 (37%) | 345 (48%) | 426 (56%) |
| **Chronic conditions <sup>d</sup>** | 150 (21%) | 161 (22%) | 163 (22%) | 159 (22%) | 166 (23%) |
| **Vaccinated** |  |  |  |  |  |
| **Overall** | 0 | 0 | 0 | 184/722 (25%) | 345/751 (46%) |
| **<12yr** |  |  |  | 0 | 93/325 (29%) |
| **≥12yr** |  |  |  | 184/345 (53%) | 252/426 (59%) |
| **Questionnaires completed** | 678 (90%) | 647 (86%) | 636 (85%) | 610 (81%) | 515 (69%) |

### 2. Repeated cross-sectional cohorts: Development of seroprevalence

Figure 2 shows the seroprevalence of children and adolescents for each timepoint of testing. Seroprevalence increased with each testing round in children and adolescents. Between T4 (Nov/Dec 2021) and T5 (Jun/Jul 2022) seroprevalence increased from 46.4% [95% credible interval [CrI] 42.6-51.0%] to 96.9% [95% CrI 95.2-98.1%] in the overall study population and from 31.3% [95% CrI 27.5-35.9%] to 95.8% [95% CrI 93.1-97.8%] among unvaccinated children and adolescents. At T4, 25.3% of all children and adolescents were vaccinated (all of whom were ≥12yrs old), and at T5 43.4% (28% of children <12 and 58% of adolescents ≥12yrs old) (see Table 1). When stratifying according to age, children below the age of 12 had a bigger increase in seroprevalence between T4 and T5 (28.4% [95% CrI 24.2-33.2%] to 95.7% [95% CrI 93.4-97.4%], respectively) compared to adolescents of 12 years or more (69.4% [95% CrI 64.0-75.4%] to 98.4% [95% CrI 97.3-99.2%], respectively) (Supplementary Table 2).

**Figure 2:**
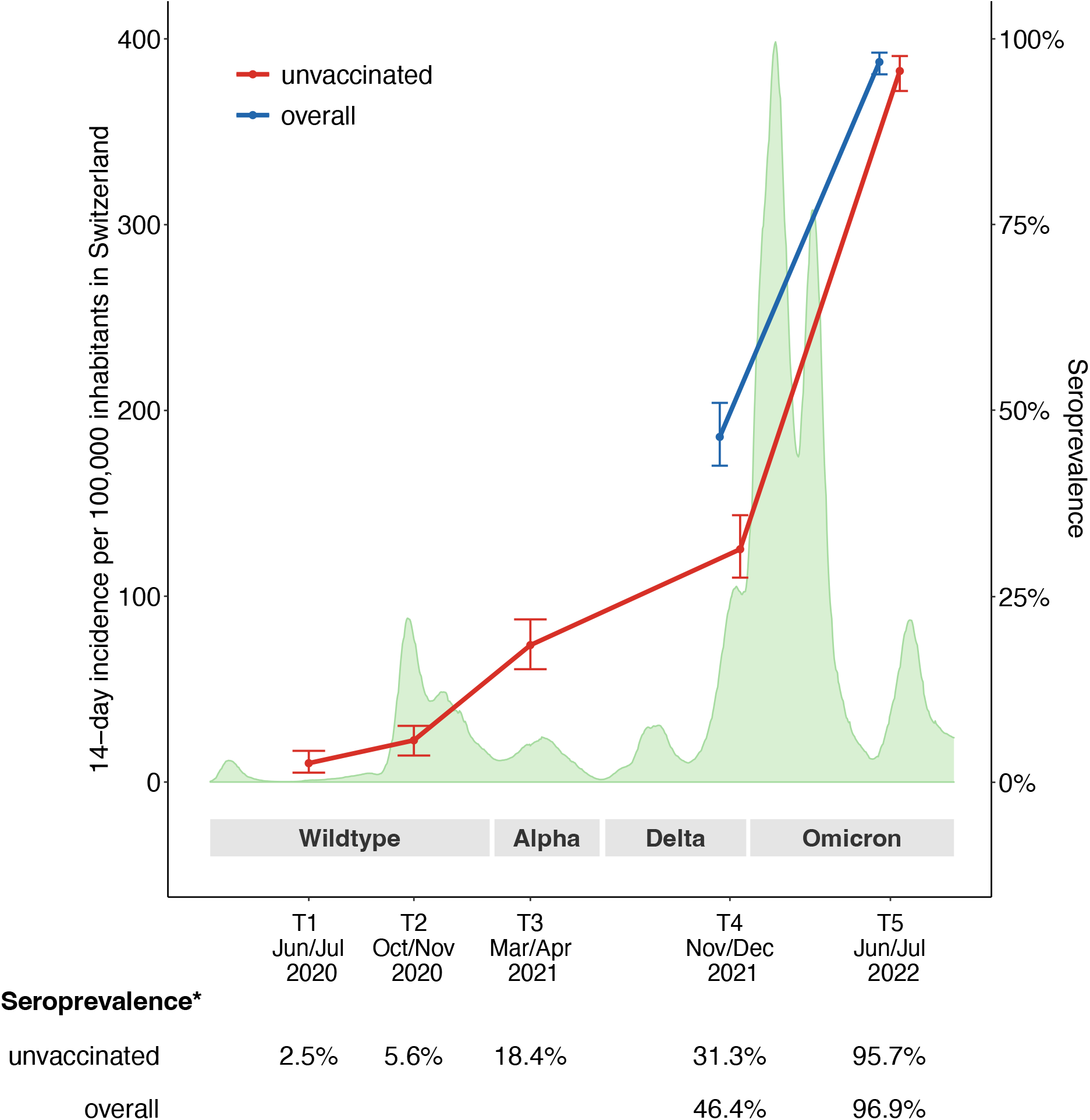
*Evolution of the incidence of diagnosed SARS-CoV-2 infections in Switzerland and participants’ seroprevalence from June 2020 to July 2022 using the cross-sectional cohort. Unvaccinated (light blue): unvaccinated children and adolescents across all 5 testing rounds; overall (dark blue): all children and adolescents participating; grey: predominant variant of concern in Switzerland (>50% of VOC circulating); *Seroprevalence was adjusted for school level, sex and district and test sensitivity and specificity*.

### 3. Longitudinal Cohort: Trajectory of Anti-spike IgG Antibodies

Figure 3 shows the trajectory of anti-spike IgG antibodies using the longitudinal cohort (n = 386) excluding children and adolescents who never tested seropositive throughout all five testing rounds (n= 37) and children and adolescents who seroconverted between T4 (Nov/Dec 2021) and T5 (Jun/Jul 2022) (n= 328). We categorised participants into four groups according to their time of seroconversion, e.g., group 1 seroconverted before T1 (Jun/Jul 2020), group 2 seroconverted between T1 (Jun/Jul 2020) and T2 (Oct/Nov 2020), group 3 seroconverted between T2 (Oct/Nov 2020) and T3 (Mar/Apr 2021), and group 4 seroconverted between T3 (Mar/Apr 2021) and T4 (Nov/Dec 2021). Anti-spike IgG antibodies remained detectable 6 months (T4 to T5: 99% (n= 208/210)), 12 months (T3 to T5: 99% (n= 113/114)), 18 months (T2 to T5: 93% (n= 28/30)) or 24 months (T1 to T5: 69% (n= 22/32)) in children and adolescents after seroconversion, respectively. At T5, antibodies were still detectable in 99% (n= 384/386) of all children and adolescents who seroconverted in any previous testing round (Supplementary Figure 1 shows the MFI ratio converted to U/ml for Roche Elecsys anti-spike IgG (WHO measure)). Anti-spike IgG titres increased with each testing round either by reinfection, vaccination, or a combination of the two. A first increase in antibody titres occurred between T3 and T4, coinciding with the introduction of vaccination in this age group in Switzerland. The highest increase in titres, visualised by the most substantial colour change in Figure 3, occurred between T4 and T5 when Omicron became the predominant VOC.

**Figure 3:**
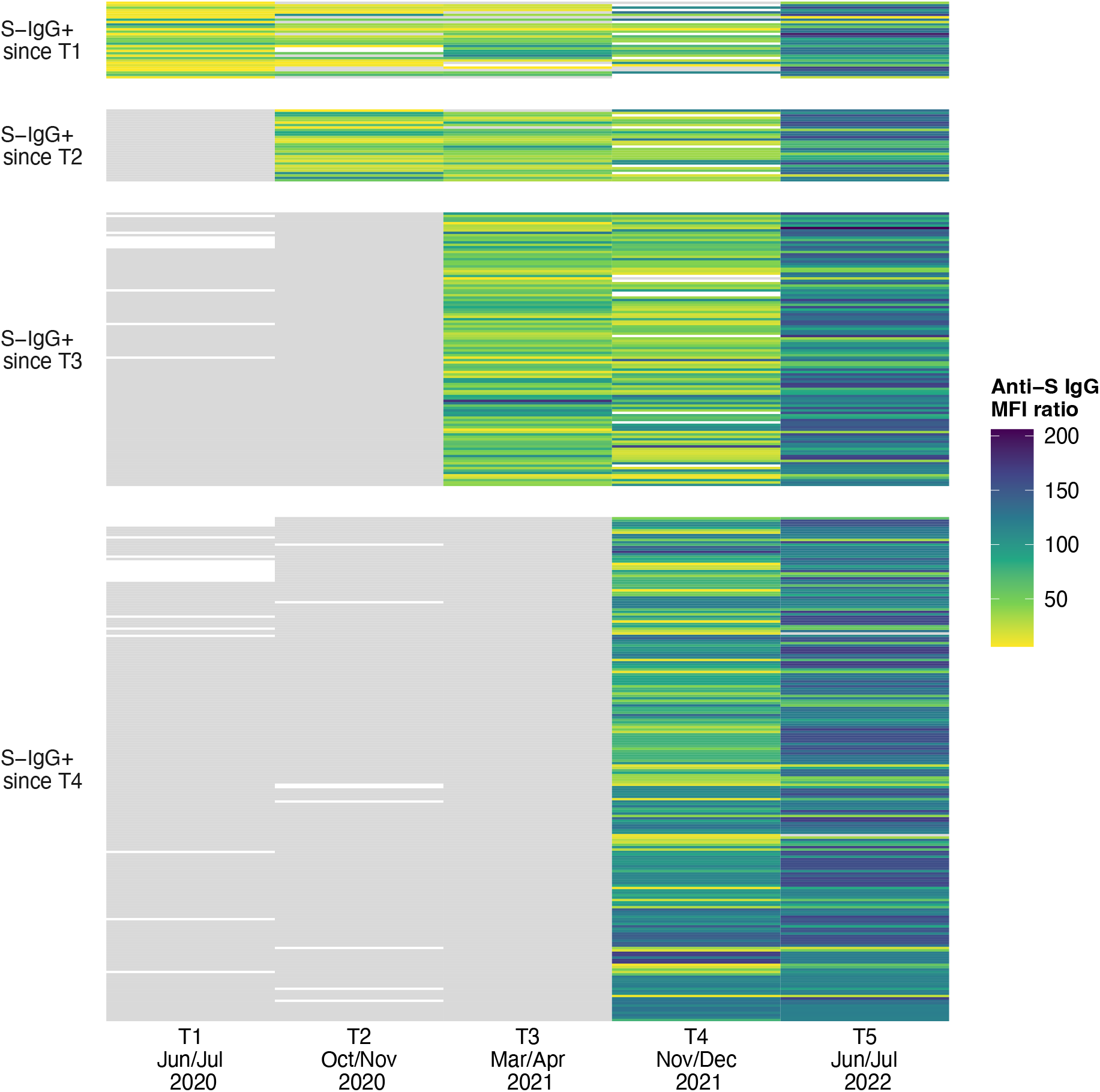
*Individual trajectories of anti-spike IgG (anti-S IgG) mean fluorescence intensity (MFI) ratios over time separated by first incidence of seropositive result (n= 386). S-IgG+: Anti-Spike IgG positive MFI ratio; 1) S-IgG+ since T1: n= 32; 2) S-IgG+ since T2: n= 30; 3) S-IgG+ since T3: n= 114; 4) S-IgG+ since T4: n= 210. Children and adolescents seroconverting from T4 to T5 are not shown (n= 328). Grey denotes seronegative anti-spike IgG result (≤6 U/ml). Colour denotes seropositive anti-spike IgG result with different level titres. White colour indicates no blood result available. 37 children and adolescent tested seronegative throughout all five testing rounds are not shown in the figure*.

To better understand the duration of protection of infection-elicited antibodies in children and adolescents, we evaluated the decay of anti-spike IgG. In a subpopulation of unvaccinated children and adolescents, excluding all children and adolescents with potential reinfection (detected by the presence of anti-nucleocapsid IgG or any increase in titres between two testing points), we estimated the anti-spike IgG antibody half-life after following them up for 365 days. The anti-spike IgG half-life estimate for this primary analysis was 305 days [95% CI 263-363 days] (Supplementary Figure 2A). For the sensitivity analysis, considering a time window of 220 days of follow up, the half-life estimate was 220 days [95% CI 170-312 days] (Supplementary Figure 2B).

### 4. Effect of the Omicron Wave

#### 4.1. **Longitudinal Cohort: Evolution of Anti-spike-IgG Antibodies**

Figure 4 shows the evolution of anti-spike IgG antibodies in groups of children and adolescents separated by their serology and exposure status (i.e., seronegative, only infected, only vaccinated, with hybrid immunity) at T4 (Nov/Dec 2021) and followed to T5 (Jun/Jul 2022). This was the time when Omicron started to be the dominant VOC until the end of the first large infection peak (Figure 2). Children and adolescents with hybrid immunity and who were only vaccinated showed similarly high titre levels at T4 or T5, whereas infected but unvaccinated children and adolescents showed considerably lower titres (Supplementary Table 3 or 4). Yet, titres of all groups increased from T4 to T5 on average. The highest increase in titres was seen in children and adolescents who were infected or seronegative in T4 and received their first vaccination between T4 and T5. The smallest increase and the lowest titres in T4 and T5 were observed in children and adolescents who were previously seronegative and had their first SARS-CoV-2 infection (Supplementary Figure 3 and Supplementary Table 4 and 5 shows the MFI ratio converted to U/ml for Roche Elecsys anti-spike IgG (WHO measure)).

**Figure 4:**
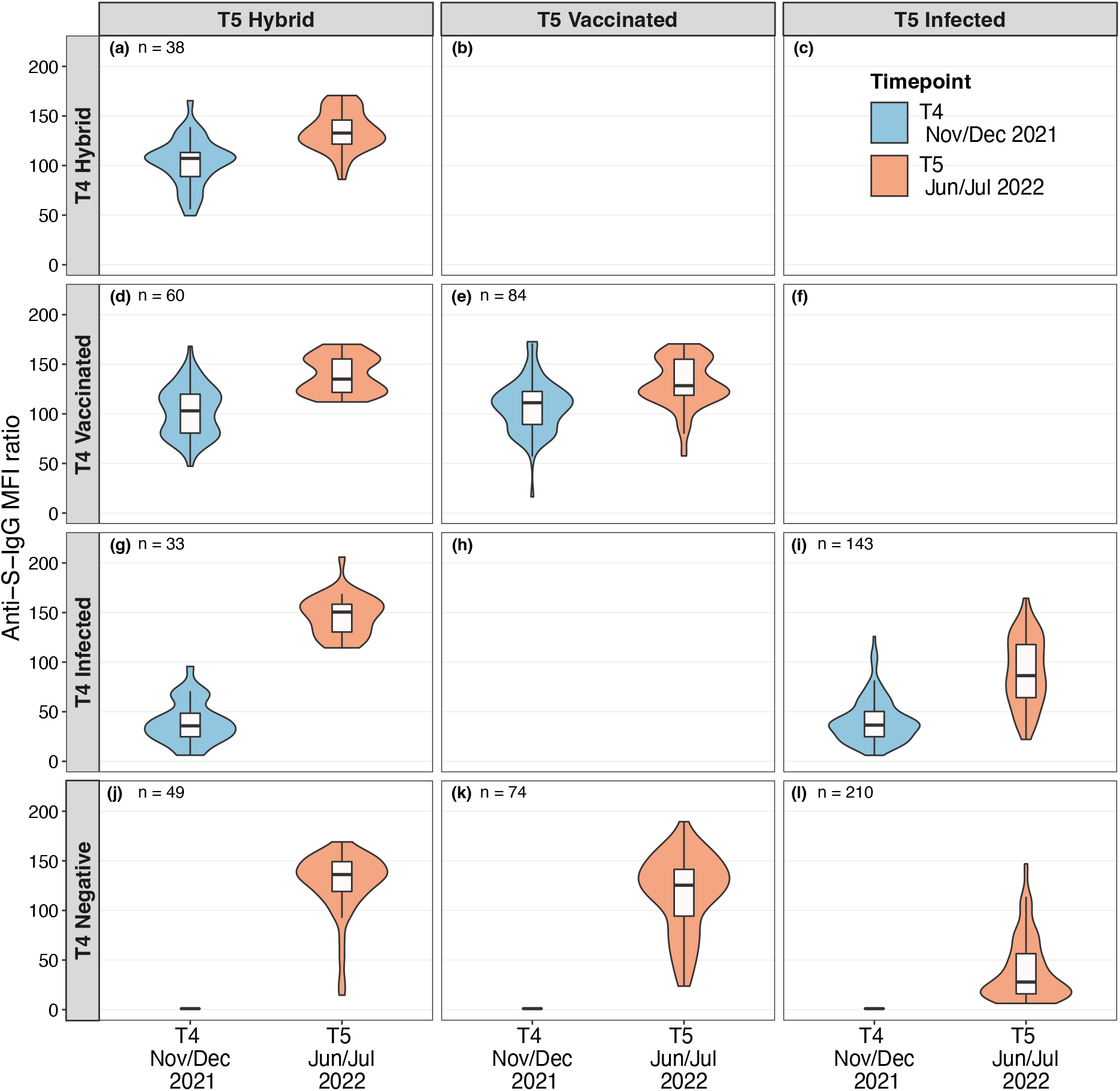
*Evolution of anti-spike IgG MFI ratios between T4 (Nov/Dec 2021) and T5 (June/July 2022) among seronegative, infected, vaccinated children and adolescents, and those with hybrid immunity at T4 shown as violin plots display mirrored density for each titre value (continuous distribution). For example, the top left panel represents 38 children and adolescents, who had median MFI titre of 107.1 in T4 (blue) and increased to median MFI titre of 133.1 in T5 (red)". Negative denotes seronegative at T4, Infected denotes seropositive but not yet vaccinated, Vaccinated denotes vaccinated participants, but negative in previous rounds and without evidence for anti-nucleocapsid IgG response. Children and adolescents with hybrid immunity were seropositive before getting vaccinated, or were vaccinated and tested positive for anti-nucleocapsid-IgG antibodies. Titre levels at T4 and at T5 are shown in blue and orange, respectively. Boxplots in panels show the median and interquartile range (IQR; whisker: 1.5 IQR). 60 children and adolescents are not shown in the figure (n=31 seronegative at T4 and T5, n=29 no data at T4)*.

To detect and quantify infections or reinfections between T4 (Nov/Dec 2021) to T5 (Jun/Jul 2022), we tested anti-nucleocapsid IgG antibodies, as they were expressed only after a SARS-CoV-2 infection and not after a vaccination. By mid 2022, more infections or reinfections happened in unvaccinated children and adolescents (74%) than in vaccinated (27%), indicating that vaccinated children and adolescents were likely better protected against an infection (Supplementary Table 6).

#### 4.2. Longitudinal Cohort: Evolution of the Neutralising Antibodies

Figure 5 shows the development of neutralising antibodies against different SARS-CoV-2 variants (Wildtype, Delta, and Omicron) between T4 (Nov/Dec 2021) and T5 (Jun/Jul 2022). We again separated participants into the four different groups according to their serology at T4 and exposure status (i.e., seronegative, only infected, only vaccinated, or with hybrid immunity). In general, neutralising activity increased in all groups between T4 and T5. The neutralising response was proportionally higher, but comparable in those with hybrid immunity and vaccination only (e.g., anti-Omicron at T5 98.9% [95% CI 96.0-99.7%] and 81.6% [95% CI 74.9-86.9%], respectively, Supplementary Table 3), but lower in infected participants (e.g., anti-Omicron at T5 64.9% [95% CI 59.8-69.7%]). The neutralising response at T5 was higher for infected children and adolescents at T4 (Figure 5i) compared to seronegative children and adolescents at T4 (Figure 5l, Supplementary Table 7 shows detailed test results). Overall, neutralising response was highest against anti-Wildtype, followed by anti-Delta and anti-Omicron, except in children and adolescent getting newly infected between T4 to T5 (Figure 5l), where anti-Omicron showed highest neutralising response, followed by anti-Wildtype and anti-Delta.

**Figure 5:**
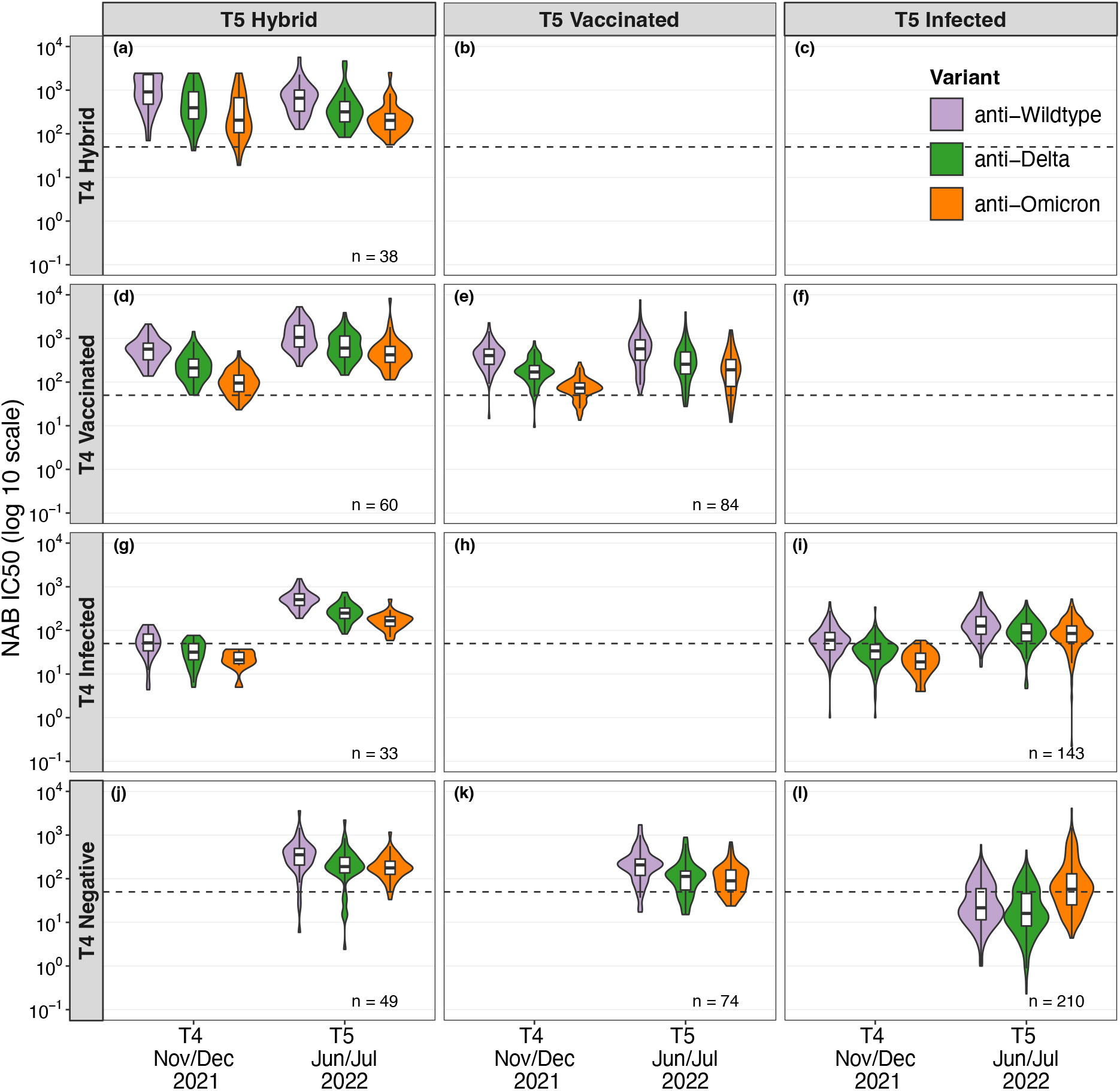
*Evolution of neutralising antibody half maximal inhibitory concentrations (IC50) between T4 (Nov/Dec 2021) and T5 (June/July 2022) among negative, infected, vaccinated children and adolescents, and those with hybrid immunity shown by violin plots display mirrored density for each NAB IC50 value (continuous distribution). Negative denotes seronegative at T4, Infected denotes seropositive but not yet vaccinated, Vaccinated denotes vaccinated participants, but negative in previous rounds and without evidence for anti-nucleocapsid IgG response. Children and adolescents with hybrid immunity were seropositive before getting vaccinated, or were vaccinated and tested positive for anti-nucleocapsid IgG antibodies. Dotted line indicates NAB IC50 value threshold (50) for half maximal inhibitory concentrations (IC_50_*) *for neutralising activity. Children and adolescents with NAB IC50 values above the threshold are assumed to have 50% or higher neutralisation capacity. Boxplots in panels show the median and interquartile range (IQR; whisker: 1.5 IQR). 60 children and adolescents are not shown in the figure (n=31 seronegative at T4 and T5, n=29 no data at T4)*.

## Discussion

Throughout the course of the Ciao Corona study from June 2020 to July 2022, seroprevalence increased with each further circulating VOC and uptake of vaccination in children and adolescents. By July 2022 despite incomplete uptake of vaccination (58% in those ≥12 years, 28% in <12 years), 96.9% [95% CrI 95.2-98.1%] of all children and adolescents had anti-spike IgG antibodies against SARS-CoV-2 and most children under the age of 12 became seropositive during the Omicron wave despite low vaccination uptake. Also, 93% of children and adolescents who seroconverted early in the pandemic were persistently seropositive for up to 18 months. Furthermore, vaccinated children and adolescents regardless of prior infection had high to very high anti-spike IgG titres and proportionally higher neutralising response, compared to unvaccinated but infected children and adolescents who showed lower SARS-CoV-2 anti-spike IgG titres and neutralising responses.

We found a sharp increase in seroprevalence from 46.4% in T4 (Nov/Dec 2021) to 96.9% in T5 (Jun/Jul 2022), and anti-spike IgG antibody titre levels in seropositive participants were overall higher at T5 compared to the beginning of the COVID-19 pandemic. While this is not surprising for those receiving a vaccination, mostly adolescents of 12 years or more, it is remarkable that also most without vaccination, i.e., children below the age of 12, became seropositive. This is likely attributable to the overall high incidence of SARS-CoV-2 infections in the first half of 2022, where the highly transmissible Omicron variant that was able to evade both natural and vaccine-induced immunity became dominant [15–17]. These findings are consistent with other international studies in children and adolescents also reporting high seroprevalence and titre levels by mid 2022 [4, 36–38]. However, seroprevalence is dependent on the course of the pandemic, the circulating VOC, geographical region, and the uptake of vaccines, which is why great variability in seroprevalence estimates were stated across different reports [39].

In our study, children and adolescents who seroconverted early in the COVID-19 pandemic, were persistently seropositive for up to 18 months. Several studies reported on duration of persistence of antibodies after a SARS-CoV-2 infection in children and adolescents. Some studies reported that anti-spike IgG levels decrease within 4 to 6 months [40, 41], whereas others showed that they remained detectable up to 9 to 18 months [9, 42, 43].

We likewise estimated that the anti-spike IgG half-life was 305 days in children and adolescents overlooking a window of 365 days. The half-life estimate of our sensitivity analysis using a time window of 220 days, was comparable to those of adults reporting between 145 to 238 days [8, 29–33]. Data on anti-spike IgG half-life in children and adolescents is limited and results are controversial, ranging from faster decay of anti-spike IgG antibodies [8] to similar [9] waning between children and adults. Thus, the estimate of half-life in our primary analysis was higher than that of the sensitivity analysis and also higher than what has previously been reported in adults. Several factors may explain these differences including the unknown timepoint of infection, differential missingness due to immune function, as well as differing assumptions made in the analyses. In particular, the sensitivity analysis with the shorter time window covered only the early time after infection in which a faster decay of antibodies takes place, while the primary analysis covered the period of a full year including both the initial fast decline of antibodies followed by a period in which the decay was much slower and steadier [8, 9].

Comparing the four groups, we found that anti-spike IgG antibody titres and neutralising response were higher in children and adolescents with hybrid immunity or vaccination only compared to only infected children and adolescents in July 2022. Neutralising capacity was proportionally higher against all VOC in children and adolescents with hybrid immunity or who were vaccinated compared to children and adolescents being only infected. We may have underestimated the proportion of children and adolescents with hybrid immunity, considering that anti-nucleocapsid IgG antibodies wane quickly and are also less expressed among vaccinated individuals [44–46]. Therefore, we likely missed infections in early 2022 if anti-nucleocapsid IgG were undetectable despite infection. However, since anti-spike IgG titres and neutralisation were similar among those vaccinated or with hybrid immunity in comparison with adults [37, 47–49], it is unlikely that this would have relevantly changed our findings. Numerous studies demonstrate that adults with high antibody titres and neutralising activity are protected against developing a severe course of SARS-CoV-2 disease [50–52]. Because only one adolescent had a hospital stay of less than 24 hours likely related to a SARS-CoV-2 infection, it remains unclear whether the findings in adults of higher protection against severe disease by vaccination and/or hybrid immunity can also be translated to children and adolescents. A study to test this hypothesis in youth would require an extremely large population-based study, as children and adolescents have a severe course of disease in less than 1 %, far less than observed in adults [53, 54].

Our findings show that the overall humoral and population immunity is high in children and adolescents in Switzerland. Policy makers and healthcare professionals in Switzerland closely followed our study and considered these findings to guide their decisions on public health recommendations and pandemic-related measures for children and adolescents. Repeated discussions of our results with cantonal and national public health authorities contributed to the decision of the Swiss Federal Office of Public Health that vaccination for youth below the age of 16 was no longer recommended, and all preventive measures regarding children and adolescents were lifted [55].

The Ciao Corona study is unique and one of few large longitudinal studies in youth [9, 37, 43]. We were able to reflect the time periods of the circulation of the major SARS-CoV-2 variants (Wildtype, Alpha, Delta, and Omicron) during our five testing rounds between June 2020 and July 2022. Also, serological testing allowed us to detect children and adolescents with asymptomatic SARS-CoV-2 infection. With the longitudinal cohort we were able to assess temporal changes in humoral activity during the COVID-19 pandemic. Furthermore, our study is the first to show the proportion of children and adolescents with neutralising antibodies based on a large school-based study.

However, some limitations need to be considered when interpreting the findings of this study. First, the exact timing of SARS-CoV-2 infections in children and adolescents is not known in sero-epidemiological studies. Thus, infection could have occurred days to months before a participant tested seropositive in our study. Second, we may have misclassified some children and adolescents when classifying them into the four different groups. Vaccination status was self-reported by the study participants or their parents. This could have led to an over- or underestimation of seroprevalence and differences in antibodies in the groups, due to recall bias. Differentiation between children and adolescents with hybrid immunity or vaccination only was based on the presence of SARS-CoV-2 anti-nucleocapsid IgG antibodies. Since the anti-nucleocapsid IgG antibodies response is weaker when vaccinated (as shown in different studies [44–46]), we likely underestimated children and adolescents with hybrid immunity. Third, the estimation of anti-spike IgG half-life bears the limitations that the decay was calculated using the first seropositive result and the time between our testing rounds varied between 4 to 8 months. Due to the missing information on the exact timepoint of infection, we set the peak at the first seropositive result to measure the decline in anti-spike IgG. However, this approach may possibly underestimate maximum antibody titers and enhancing variability of measured values. Consequently, we may have overestimated the half-life in children and adolescents. Fourth, persistence of anti-spike IgG antibodies over 24 months in 69% of children and adolescents may be underestimated due to false positive serological results at T1 (Jun/Jul 2020) when SARS-CoV-2 prevalence was low [56].

## Conclusion

In this study, we highlighted the importance of serological studies as a COVID-19 monitoring tool and the development of humoral immunity in children and adolescents. Our findings show that the Omicron wave and the rollout of vaccines led to almost 100% seropositivity and boosted seroprevalence and anti-spike IgG antibody titres (by infection, reinfection, and/or vaccination) as well as led to better neutralising capacity in children and adolescents. Especially during the first peak of the Omicron wave, most unvaccinated children under the age of 12 became seropositive compared to adolescents of 12 years and older, who had access to vaccines since June 2021. Nevertheless, during the entire study period parents of three children and adolescents reported a hospital stay of less than 24 hours, of which one was possibly related to an acute SARS-CoV-2 infection.

## Authors Contributions

SK and MAP came up with the study idea. SK, MAP, TR, and JF developed the preliminary design. SK, MAP, TR and AU established the study design and methodology. SK, AU, TR, SR, SRH and AR performed participant recruitment, data collection and management. SRH, SR, AU and AR conducted the data cleaning and the statistical analysis. GP, CF, CP and DLC devised the serology analysis protocol and supervised, conducted, and assessed the serological examinations. AR wrote the first draft of the manuscript. All authors were involved in the interpretation of the findings, the review and authorisation of the manuscript for intellectual accuracy. SK is the corresponding author and guarantor, assuming complete accountability for the conducted research. Furthermore, SK had full access to the data, and made the final decision to publish. The corresponding author (SK) attests that all listed authors meet authorship criteria and that no others meeting the criteria have been omitted.

## Ethics approval

The Ethics Committee of the Canton Zurich, Switzerland, approved the study (2020-01336). All study participants provided written informed consent prior to study enrolment.

## Transparency declaration

The lead authors affirm that the manuscript is an honest, accurate, and transparent account of the study being reported, no important aspects of the study have been omitted, and any discrepancies from the study as originally planned and registered have been explained.

### Copyright

The Corresponding Author has the right to grant on behalf of all authors and does grant on behalf of all authors, a worldwide licence to the Publishers and its licensees in perpetuity, in all forms, formats and media (whether known now or created in the future), to i) publish, reproduce, distribute, display and store the Contribution, ii) translate the Contribution into other languages, create adaptations, reprints, include within collections and create summaries, extracts and/or, abstracts of the Contribution, iii) create any other derivative work(s) based on the Contribution, iv) to exploit all subsidiary rights in the Contribution, v) the inclusion of electronic links from the Contribution to third party material where-ever it may be located; and, vi) licence any third party to do any or all of the above.

## Data Availability

All data produced in the present study are available upon reasonable request to the authors.

