## Supplementary for "Persistent humoral immunity in children and adolescents throughout the COVID-19 pandemic (June 2020 to July 2022): a prospective school-based cohort study (Ciao Corona) in Switzerland"

### Supplementary Material

**Supplementary Table 1: Chronic condition in children and adolescents.** Chronic disease assessed via questionnaires. In this table are listed all the chronic conditions reported by the children and adolescents or their proxy.

| |
|---|
| Asthma |
| hay fever |
| celiac disease |
| lactose intolerance |
| allergies (other than hay fever) |
| neurodermitis |
| diabetes mellitus |
| inflammatory bowel disease |
| hypertension |
| arthritis |
| other chronic diseases potentially affecting the immune response (Neutropenia, PFAPA-Syndrome, renal failure, cystic fibrosis, bronchitis) |

**Supplementary Table 2: Overall seroprevalence and seroprevalence split in ≥12 and <12-year-old children and adolescents.** This table illustrates the overall seroprevalence and seroprevalence divided according to age (<12 years and ≥ 12years) for all five testing rounds. The proportion of vaccinated children and adolescents per testing round is also indicated.

| | **Timeframe of testing** | **T1**<br>**Jun/July 2020** | **T2**<br>**Nov/Dec 2020** | **T3**<br>**Mar/Apr 2021** | **T4**<br>**Oct/Nov 2021** | **T5**<br>**Jun/Jul 2022** |
|---|---|---|---|---|---|---|
| **All** | **Seroprevalence:**<br><br>Unvaccinated | 3.0% | 5.6% | 18.4% | 31.3% | 95.8% |

| | | | | | | |
|---|---|---|---|---|---|---|
| | | (1.3% - 4.5%) | (3.6% - 7.6%) | (15.2% - 21.9%) | (27.5% - 35.9%) | (93.1% - 97.8%) |
| | Vaccinated ± infected | - | - | - | 46.4%<br>(42.6% - 51.0%) | 96.9%<br>(95.2% - 98.1%) |
| | % vaccinated | - | - | - | 25.5% | 43.4% |
| **<12yrs** | **Seroprevalence:** | | | | | |
| | Unvaccinated | 3.0%<br>(1.4% - 4.6%) | 5.6%<br>(3.5% - 7.8%) | 19.7%<br>(16.0% - 24.0%) | 28.4%<br>(24.2% - 33.2%) | 95.1%<br>(92.2% - 97.5%) |
| | Vaccinated ± infected | - | - | - | - | 95.7%<br>(93.4% - 97.4%) |
| | % vaccinated | - | - | - | 0 | 28.0 % |
| **≥12yrs** | **Seroprevalence:** | | | | | |
| | Unvaccinated | 2.9%<br>(0.9% -4.9%) | 5.6%<br>(3.2% - 7.9%) | 16.8%<br>(13.5% - 20.1%) | 40.3%<br>(34.2% - 47.0%) | 97.2%<br>(94.7% - 98.7%) |
| | Vaccinated ± infected | - | - | - | 69.4%<br>(64.0% - 75.4%) | 98.4%<br>(97.3% - 99.2%) |
| | % vaccinated | - | - | - | 51.2% | 57.6% |

**Supplementary Table 3: Anti-spike IgG MFI ratios and neutralising activity (IC50) at T5.** This table provides detailed anti-spike IgG titres (MFI ratio, positive values defined by a cutoff of 6 or higher) and neutralising activity (measured by the half maximal inhibitory concentration IC50, with positive values defined by a cutoff of 50 or higher) in children and adolescents from the longitudinal cohort. We divided individuals based on their serology at T5 in Hybrid, Vaccinated and Infected.

| N | T5 Group | Anti-spike IgG titres (MFI Ratio) Median and IQR | Anti-Wildtype neutralising activity (IC50) Median (IQR) | Anti-Delta neutralising activity (IC50) Median (IQR) | Anti-Omicron neutralising activity (IC50) Median (IQR) | % of Individuals above threshold for Anti-Wildtype (95% CI) | % of Individuals above threshold for Anti-Delta (95% CI) | % of Individuals above threshold for Anti-Omicron (95% CI) |
|---|---|---|---|---|---|---|---|---|
| 180 | Hybrid | 136.2 (121.9-154.3) | 591.2 (355.4-1055.2) | 316.9 (189.1-574.1) | 220 (147.7-382.9) | 98.3 (95.2-99.4) | 96.1 (92.2-98.1) | 98.9 (96.0-99.7) |
| 158 | Vaccinated | 127.6 (114.1-151) | 312.8 (173-689.4) | 158.4 (88.9-337.4) | 133.2 (60.5-214.4) | 96.2 (92.0-98.2) | 86.7 (80.5-91.1) | 81.6 (74.9-86.9) |
| 353 | Infected | 54.8 (22.8-89.8) | 59.2 (16.7-125.9) | 42.5 (11.9-88.3) | 69.5 (34.2-127.4) | 54.4 (49.2-59.5) | 45.6 (40.5-50.8) | 64.9 (59.8-69.7) |

**Supplementary Table 4: Anti-spike IgG MFI ratios between T4 (Nov/Dec 2021) and T5 (Jun/Jul 2022).** This table provides the anti-spike IgG MFI ratio (median and interquartile range) shown in the boxplot (Figure 4) for all different group combinations.

| N | T4 Group | T5 Group | T4: Median (IQR) MFI ratio | T5: Median (IQR) MFI ratio |
|---|---|---|---|---|
| **38** | T4 Hybrid | T5 Hybrid | 107.2 (88.8-113.1) | 132.8 (121.6-145.9) |
| **60** | T4 Vaccinated | T5 Hybrid | 103 (80.6-119.8) | 135.1 (121.6-155.3) |
| **84** | T4 Vaccinated | T5 Vaccinated | 111.2 (89.3-122.6) | 128.4 (118.7-155.1) |
| **33** | T4 Infected | T5 Hybrid | 35.8 (24.8-48.5) | 150.5 (130.4-158.4) |
| **143** | T4 Infected | T5 Infected | 36.5 (24.9-50.2) | 86.4 (64.2-117.8) |
| **49** | T4 Negative | T5 Hybrid | 1 (1-1) | 136.2 (119.1-149.2) |
| **74** | T4 Negative | T5 Vaccinated | 1 (1-1) | 125.4 (94.3-141.4) |
| **210** | T4 Negative | T5 Infected | 1 (1-1.2) | 27.9 (15.9-56.4) |

**Supplementary Table 5: Anti-spike IgG MFI ratio converted to U/ml for Roche Elecsys anti-spike IgG (WHO measure) between T4 (Nov/Dec 2021) and T5 (Jun/Jul 2022).** This table provides the MFI ratio converted to U/ml for Roche Elecsys anti-spike IgG (WHO measure) shown in the boxplot (Supplementary Figure 3). We converted the MFI values to U/ml, by using the Elecsys Anti-SARS-CoV2 immunoassay developed by Roche, for the purpose of interpretation. The Department of Clinical Immunology & Allergy of the University Hospital of Lausanne used population based samples to provide the conversion formula of Roche anti-spike IgG = 10^(−0.6108069 + 2.0072882 × log10(MFI + 1)).

| N | T4 Group | T5 Group | T4: Median (IQR) U/ml | T5: Median (IQR) U/ml |
|---|---|---|---|---|
| **38** | T4 Hybrid | T5 Hybrid | 2968.1 (2044.4-3303.3) | 4542.3 (3812.9-5479.4) |
| **60** | T4 Vaccinated | T5 Hybrid | 2744 (1685.8-3706.2) | 4700.7 (3815.7-6212.1) |
| **84** | T4 Vaccinated | T5 Vaccinated | 3192.4 (2064.5-3875.3) | 4250.6 (3636.7-6190.2) |
| **33** | T4 Infected | T5 Hybrid | 340.6 (167-617.7) | 5833.2 (4383.5-6459.8) |
| **143** | T4 Infected | T5 Infected | 353.8 (167.7-662.3) | 1933.6 (1075.4-3577.5) |
| **49** | T4 Negative | T5 Hybrid | 1 (1-1) | 4780.6 (3659.6-5733.2) |
| **74** | T4 Negative | T5 Vaccinated | 1 (1-1) | 4058.9 (2303.2-5151.2) |
| **210** | T4 Negative | T5 Infected | 1 (1-1.2) | 209 (71.6-832.2) |

**Supplementary Table 6: Proportion of children and adolescents with reinfection between T4 (Nov/Dec 2021) and T5 (Jun/Jul 2022).** This table shows the proportion of children and adolescents with first SARS-CoV-2 infection before T4 (Nov/Dec 2021) and reinfection between T4 (Nov/Dec 2021) and T5 (Jun/Jul 2022). We divided children and adolescents into unvaccinated, recent vaccination (latest vaccination between T4 and T5) and older infection (latest vaccination before T4). In these three groups we calculated the proportion of children and adolescents experiencing reinfection by looking at the presence of anti-nucleocapsid IgG as well as 25% increase and/or decrease.

| | **Unvaccinated** N=314 | | **Recent vaccination** N=220 | | **Older vaccination** N=206 | |
|---|---|---|---|---|---|---|
| Anti-nucleocapsid IgG positivity | Anti-N IgG + | Anti-N IgG - | Anti-N IgG + | Anti-N IgG - | Anti-N IgG + | Anti-N IgG - |
| N ≥25% titre increase | 231/314 (73.6%) | 31/314 (9.9%) | 57/220 (25.9%) | 100/220 (45.5%) | 58/206 (28.2%) | 53/206 (25.7%) |

| | | | | | | |
|---|---|---|---|---|---|---|
| N between <25% titre increase & <25% titre decrease | 17/314 (5.4%) | 23/314 (7.3%) | 23/220 (10.5%) | 34/220 (15.5%) | 42/206 (20.4%) | 40/206 19.4% |
| N ≥25% titre decrease | 4/314 (1.3%) | 8/314 (2.5) | 1/220 (0.4%) | 5/220 (2.3%) | 2/206 (0.9%) | 11/206 (5.4%) |

**Supplementary Table 7: Neutralising activity between T4 (Nov/Dec 2021) and T5 (Jun/Jul 2022).** This figure shows the development of neutralising antibodies against different SARS-CoV-2 variants between T4 (Nov/Dec 2021) and T5 (Jun/Jul 2022). We again separated participants into the four different groups according to their serology at T4 and exposure status (i.e., seronegative, only infected, only vaccinated, or with hybrid immunity) and evaluated neutralising activity against Wildtype, Delta, and Omicron. We calculated medians (with inter quartile ranges (IQR)) and proportion of children and adolescents with neutralising activity, measured by the half maximal inhibitory concentration (IC50) with positive or negative results defined by a cutoff value of 50 or higher.

| **N** | **Variant of concern** | **T4 Group** | **T5 Group** | **T4: Median (IQR)** | **T5: Median (IQR)** | **T4: % of Individuals above threshold (%)** | **T5: % of Individuals above threshold (%)** |
|---|---|---|---|---|---|---|---|
| 38 | Wildtype | T4 Hybrid | T5 Hybrid | 913.9 (476.9-2287.4) | 672.5 (339.1-1015.2) | 100 (90.8-100) | 100 (90.8-100) |
| 60 | Wildtype | T4 Vaccinated | T5 Hybrid | 568.5 (324.4-780.8) | 1082.9 (639.9-2083.3) | 100 (94.0-100) | 100 (94.0-100) |
| 84 | Wildtype | T4 Vaccinated | T5 Vaccinated | 405.2 (254.9-569.3) | 578.1 (319.1-929.3) | 98.8 (93.6-99.8) | 100 (95.6-100) |
| 33 | Wildtype | T4 Infected | T5 Hybrid | 51.9 (34-82.2) | 505.8 (372.9-687.8) | 51.5 (35.2-67.5) | 100 (90.0-100) |

| | | | | | | | |
|---|---|---|---|---|---|---|---|
| 143 | Wildtype | T4 Infected | T5 Infected | 59.7 (35.8-89.4) | 125.9 (81.6-206.3) | 58 (49.8-65.8) | 90.9 (85.1-94.6) |
| 49 | Wildtype | T4 Negative | T5 Hybrid | 0 | 354 (205.4-494.1) | 0 (0-7.3) | 93.9 (83.5-97.9) |
| 74 | Wildtype | T4 Negative | T5 Vaccinated | 0 | 207.3 (119-281.8) | 0 (0-4.9) | 91.9 (83.4-96.2) |
| 210 | Wildtype | T4 Negative | T5 Infected | 0 | 21.5 (11.1-59.2) | 0 (0-1.8) | 29.5 (23.8-36.0) |
| 38 | Delta | T4 Hybrid | T5 Hybrid | 395.3 (219.4-919.9) | 318.6 (187.4-547.7) | 97.4 (86.5-99.5) | 100 (90.8-100) |
| 60 | Delta | T4 Vaccinated | T5 Hybrid | 211.3 (129.2-337.5) | 610.2 (377.7-1143.4) | 100 (94.0-100) | 100 (94.0-100) |
| 84 | Delta | T4 Vaccinated | T5 Vaccinated | 170.5 (118.4-240.4) | 256.9 (153-490.5) | 96.4 (90.0-98.8) | 94.0 (86.8-97.4) |
| 33 | Delta | T4 Infected | T5 Hybrid | 31.7 (21.2-49.7) | 249 (187.4-324.7) | 24.2 (12.8-41.0) | 100 (90.0-100) |
| 143 | Delta | T4 Infected | T5 Infected | 33.9 (22-47.7) | 88.3 (56.6-141.3) | 23.1 (16.9-30.6) | 79.7 (72.4-85.5) |
| 49 | Delta | T4 Negative | T5 Hybrid | 0 | 189.8 (136.1-308.7) | 0 (0-7.3) | 85.7 (73.3-92.9) |
| 74 | Delta | T4 Negative | T5 Vaccinated | 0 | 112.8 (55.2-150.1) | 0 (0-4.9) | 78.4 (67.8-86.2) |
| 210 | Delta | T4 Negative | T5 Infected | 0 | 16 (8.3-45.8) | 0 (0-1.8) | 22.4 (17.3-28.5) |
| 38 | Omicron | T4 Hybrid | T5 Hybrid | 191 (105.2-630.5) | 203.3 (124.7-288.8) | 92.1 (79.2-97.3) | 100 (90.8-100) |
| 60 | Omicron | T4 Vaccinated | T5 Hybrid | 95 (60.5-143.9) | 421.9 (284.1-656.5) | 81.7 (70.1-89.4) | 100 (94.0-100) |
| 84 | Omicron | T4 Vaccinated | T5 Vaccinated | 70.1 (49.8-95) | 191.7 (79.5-329.9) | 75 (64.8-83.0) | 84.5 (75.3-90.7) |
| 33 | Omicron | T4 Infected | T5 Hybrid | 0 | 166.2 (124.8-206.1) | 0 (0-10.4) | 100 (90.0-100) |
| 143 | Omicron | T4 Infected | T5 Infected | 0 (0-13) | 85.2 (54.2-126.8) | 1.4 (0.4-5.0) | 78.3 (70.9-84.3) |
| 49 | Omicron | T4 Negative | T5 Hybrid | 0 | 176.9 (125.9-252.1) | 0 (0-7.3) | 95.9 (86.3-98.9) |
| 74 | Omicron | T4 Negative | T5 Vaccinated | 0 | 89.6 (54.4-158.5) | 0 (0-4.9) | 78.4 (67.7-86.3) |
| 210 | Omicron | T4 Negative | T5 Infected | 0 | 57.3 (25.1-129.5) | 0 (0-1.8) | 55.7 (49.0-62.3) |

**Supplementary Figure 1: Heatmap for the longitudinal trajectory of MFI ratio converted to U/ml for Roche Elecsys anti-spike IgG (WHO measure) in children and adolescents over the entire study period.** Individual trajectories of anti-spike IgG of MFI ratio converted to U/ml for Roche Elecsys anti-spike IgG (WHO measure) over time separated by first incidence of seropositive result in children and adolescents (total n = 386). We excluded children and adolescents who never tested seropositive throughout all five testing rounds (n= 37) and children and adolescents who seroconverted between T4 (Nov/Dec 2021) and T5 (Jun/Jul 2022) (n= 328). We converted the MFI values to U/ml, by using the Elecsys Anti-SARS-CoV2 immunoassay developed by Roche, for the purpose of interpretation. The Department of Clinical Immunology & Allergy of the University Hospital of Lausanne used population based samples to provide the conversion formula Roche anti-spike IgG = $10^{\wedge}(-0.6108069 + 2.0072882 \times \log10(MFI + 1))$.

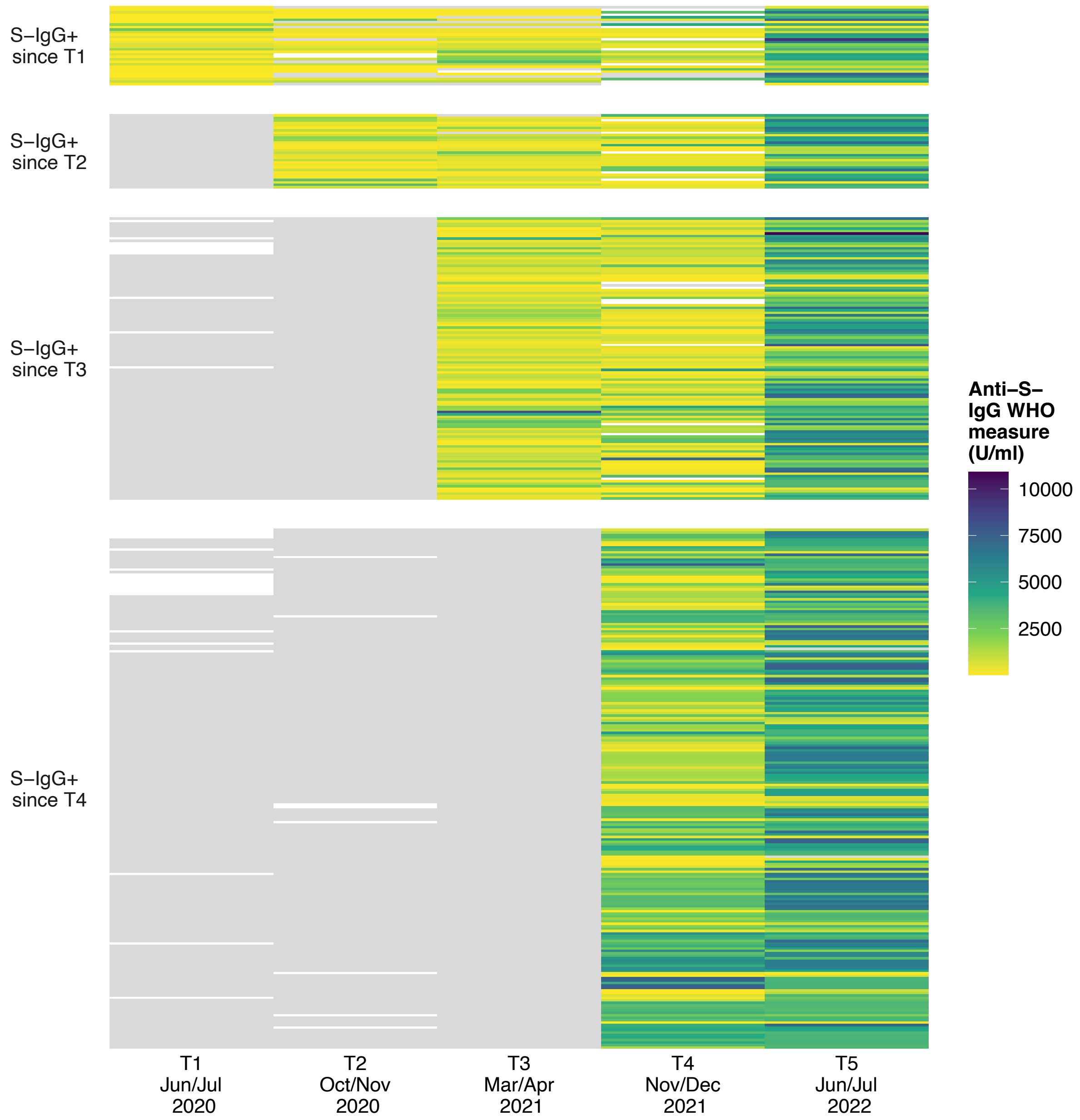


**Supplementary Figure 2: Half-life of anti-spike IgG antibodies.** For the primary analysis, we calculated the waning of anti-spike IgG antibodies in children and adolescents considering a time window of follow up of 365 days (total n= 114) (**A**). For the sensitivity analysis we used a shorter time window of 220 days (total n= 53) (**B**). We followed up all children and adolescents from the timepoint of seroconversion. We excluded individuals who never tested seropositive for anti-spike IgG antibody, and who had no follow-up assessment after being tested seropositive and those who got vaccinated. A mixed linear model was conducted to estimate the anti-spike IgG decay over time using a random intercept for participants.

**(A)**

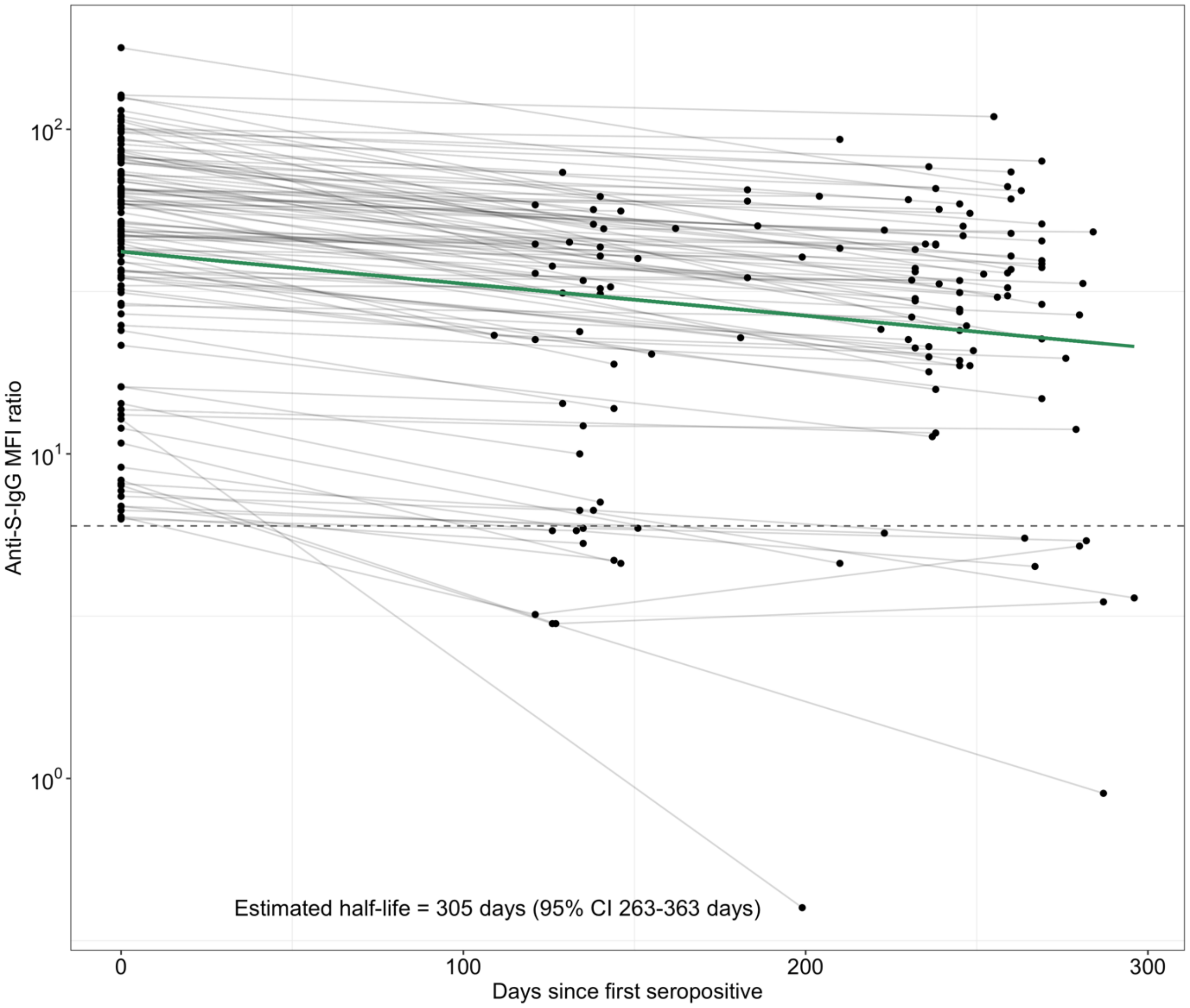


**(B)**

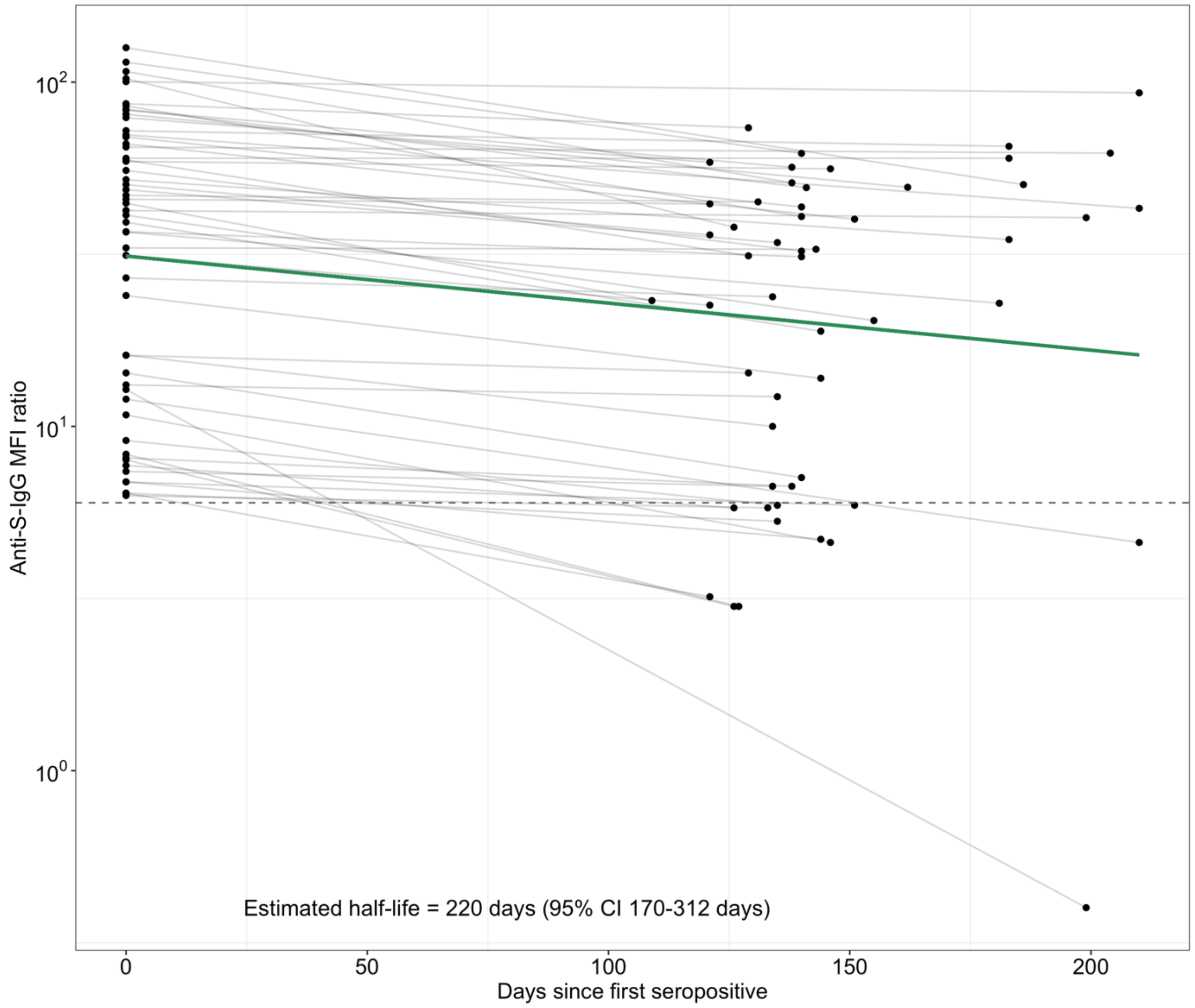


**Supplementary Figure 3: Evolution of anti-spike IgG of MFI ratio converted to U/ml for Roche Elecsys anti-spike IgG (WHO measure) from T4 (Nov/Dec 2021) to T5 (Jun/Jul 2022).** This figure shows the evolution of anti-spike IgG antibodies in groups of children and adolescents separated by their serology and exposure status (i.e., seronegative, only infected, only vaccinated, with hybrid immunity) at T4 (Nov/Dec 2021) and followed to T5 (Jun/Jul 2022). We converted the MFI values to U/ml, by using the Elecsys Anti-SARS-CoV2 immunoassay developed by Roche, for the purpose of interpretation. The Department of Clinical Immunology & Allergy of the University Hospital of Lausanne used population based samples to provide the conversion formula Roche anti-spike IgG = 10^(−0.6108069 + 2.0072882 × log10(MFI + 1)).

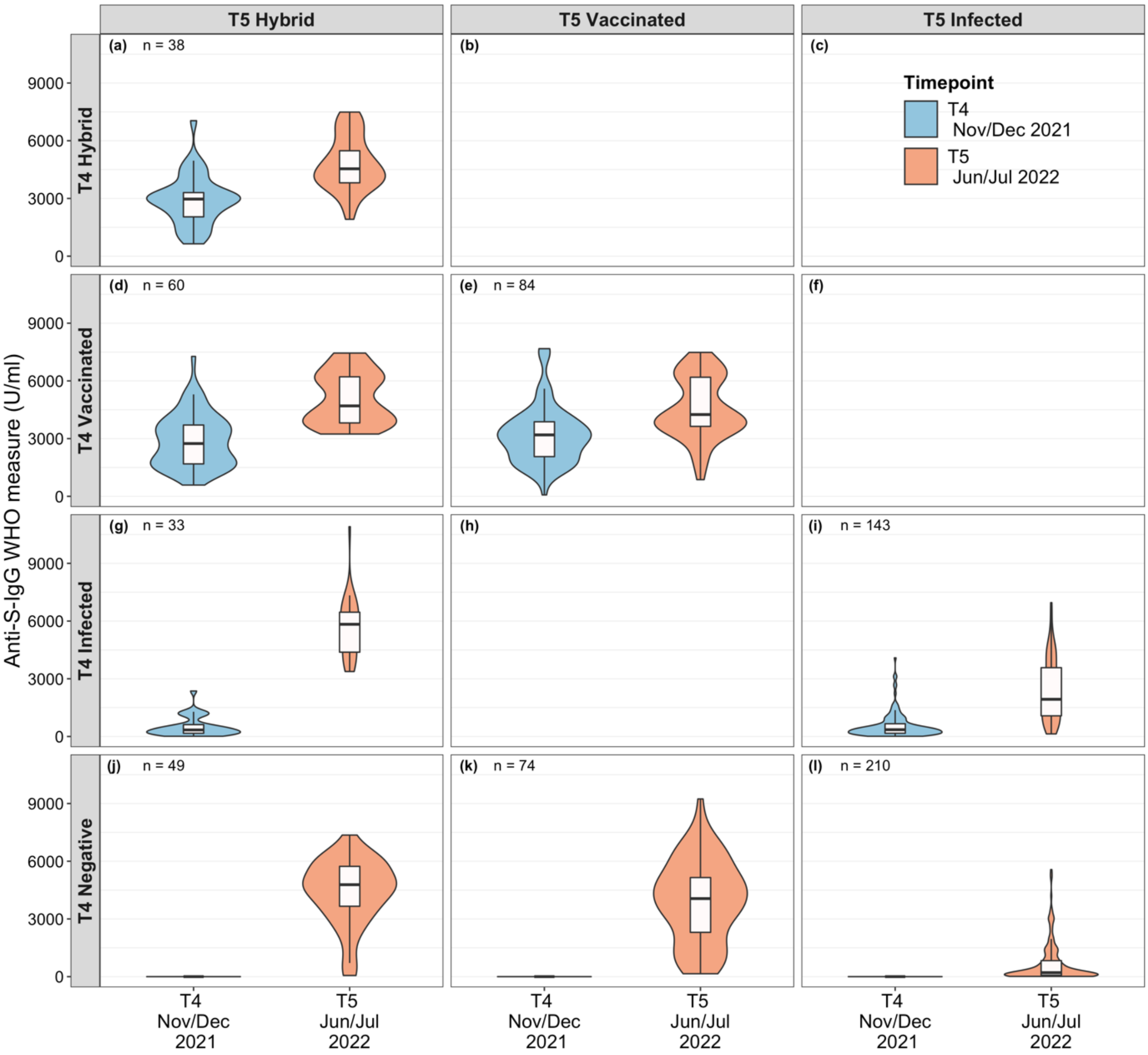
